# IVUS Improves Outcomes with SUPERA Stents for the Treatment of Superficial Femoral-popliteal Artery Disease

**DOI:** 10.1101/2023.02.03.23285427

**Authors:** Prakash Krishnan, Raman Sharma, Sriya Avadhani, Arthur Tarricone, Allen Gee, Serdar Farhan, Haroon Kamran, Annapoorna Kini, Samin Sharma

## Abstract

**Objectives:** To determine the effect of intravenous ultrasound (IVUS) when used adjunctively with nitinol interwoven bare metal stents in the management of femoropopliteal lesions.

**Background:** Nitinol interwoven bare metal stents represent an advancement in stent technology; however nominal deployment remains an area of focus. Intravascular Ultrasound (IVUS) has been shown to improve outcomes in both the coronary and peripheral vasculature by providing the operator with greater vessel detail. The use of adjunctive IVUS with Nitinol bare metal stents has not been widely studied.

**Methods:** This study included a cohort of 200 consecutive patients with peripheral artery disease. All patients were treated with ≥1 Supera bare metal stent, and 91 received adjunctive IVUS imaging prior to stent deployment. Deployment conditions of nominal, compressed, and elongated were measured and the primary clinical outcomes included target lesion reintervention, major and minor, below the knee amputation, and mortality.

**Results:** The patients who received IVUS had a significantly greater number of nominally deployed stents (p<0.001). Patients who had IVUS imaging also had significantly lower reintervention rates compared to those who did not receive IVUS imaging (p=0.047).

**Conclusion:** IVUS and angiography decreases clinically driven target lesion reintervention and increases nominal deployment compared to angiography alone in femoropopliteal lesions treated with interwoven bare metal nitinol stents.

**Condensed Abstract:** This retrospective study was done to examine the utility of intravenous ultrasound when used adjunctively for visualization in the management of femoropopliteal lesions in peripheral artery disease patients. There was total of 200 subjects and all were treated using the Supera nitinol self-expanding stent. By the end of the study, the group of subjects treated with intravenous ultrasound had significantly fewer reinterventions and had a greater rate of nominally deployed stents. These findings suggest the beneficial role intravenous ultrasound has compared to angiography alone in the management of femoropopliteal lesions with Supera stents.

## Introduction

Peripheral Artery Disease (PAD) affects over 200 million people globally with increasing prevalence among the elderly (>80 years) population.(1) Endovascular management of PAD has become the mainstay for treatment; however long term patency remains an area of focus.

Endovascular intervention success is multifactorial, with outcomes influenced by multiple variables ranging from clinical presentation to appropriate interpretation of angiography.(2,3) Stent malposition due to over or under sizing has been observed to result in increased clinically driven target lesion reintervention (CD-TLR) and decrease in primary patency. Intravascular Ultrasound (IVUS) has been shown to improve outcomes in both the coronary and peripheral vasculature by providing the operator with greater vessel detail specifically assessment of plaque buildup, measurement of lumen diameter, and possible assurance of proper stent deployment.(4) In the peripheral vasculature, IVUS use has been observed to significantly improve primary and secondary patency, freedom from any reintervention, freedom from any adverse limb event, protective against vascular and non-vascular complications, and increase event-free survival compared to angiography alone.(5-7)

The interwoven nitinol stent (Supera) represents an advancement in stenting technology and has been shown to improve primary patency compared to bare metal stents, with rates published as high as 90%.(8-11) Logical explanation of this improvement may be attributed to the inherent helical structure of Supera, allowing greater resistance to compressive arterial forces and resilience to stent fractures.(12) However, there has been variation in the success of studies, with patency rates of Supera ranging from 55% -90%.(13,14) One factor that have been speculated attributing to this variation is the mal-deployment of the Supera stents; and with fewer nominally deployed stents, there will likely be greater target lesion revascularization.(15) While nominal deployment is always the operator’s objective, success of this has only been observed in approximately 50% of treated subjects. The adjunctive use of IVUS with Supera stent technology has not been widely studied and the objective of this analysis is to examine a cohort of femoropopliteal PAD patients treated with Supera and IVUS vs Supera alone to observe differences in both deployment condition as well as clinical outcomes.

## Methods

Between March 2010 to March 2020, 200 consecutive patients were treated for femoropopliteal PAD with Supera stents. All patients were adults (age ≥18 years), had a confirmed clinical diagnosis of PAD, and were treated with at least one (≥1) Supera stent. All patients were treated by guideline directed medical therapy. This study was approved by the Institutional Review Board of the Icahn School of Medicine at the Mount Sinai Hospital: Program of Protection of Human Subjects Committee.

The utility of adjunctive procedures and the duration of dual antiplatelet therapy was under the discretion of the treating physician. Standard institutional practice of clopidogrel (75mg/day), and aspirin (81mg) for 3 months were followed for post-procedure therapy. Deployment conditions were assessed based on angiogram review and conditions were classified as nominal, compressed, and elongated by two physicians independently. All disagreements were resolved by the corresponding author

All angiograms were anonymized prior to review by the data collection team. Reference vessel diameter was established through angiography or IVUS analysis when available. The estimation of compression and elongation was determined by the operator, by dividing the nominal stent length according to package insertion directions. Actual deployment length was measured using a preprocedural attached measuring scale. Optical evaluation of the complete stent was then conducted to examine for signs of elongation or compression. In keeping with the findings in the SUPERB trial^7^, the authors defined the final three Supera stent dimensions as elongation (≥10% of package insert length), compression (≤10% of package insert length), and nominal (−10% ≤ Length ≤+10% of package insert length). 10% was chosen due to the increase in CD-TLR observed in the SUPERB trial^7^ when the Supera deployment was either compressed or elongated greater than 10%.

IVUS was performed using a system (Philips Healthcare) for all subsequent cases. Images were acquired with a Volcano Eagle Eye(0.014-inch guidewire) or PV018 (0.018in guide wire) solid-state phased array IVUS catheters. All the treating operators were experienced in the use and interpretation of IVUS and IVUS was performed after pre-dilation and prior to stent deployment. In cases with chronic total occlusion, pretreatment IVUS imaging was performed after crossing of the occlusion. Manual pull back performed at a steady rate under fluoroscopic imaging with bookmarking of frames at regular distances in relation to the radiopaque ruler for all cases. In addition for all cases, the frame rate was set to maximum and gain and field of view were optimized. Pullback acquisitions and saved still images were archived on softlink international and saved for later analysis. IVUS diameter measurements were obtained at the time of the procedure using the control module measurement software. IVUS diameters of the proximal and distal Reference Vessel Diameter (RVD) were obtained at the most normal appearing section immediately adjacent to the lesion. The IVUS diameter was measured in millimeters and obtained following institutional guidelines by measuring the distance between the intima-lumen boundaries in orthogonal planes and calculating the mean measurement. The larger of the two measurements was used as the IVUS RVD. IVUS lesion length (mm) was estimated by identifying the proximal and distal edge of the lesion according to IVUS while under direct fluoroscopic control, and the distance between these points was measured using the angiographic imaging length measurement technique described previously. Additional information including the presence of post-angioplasty dissection was also obtained.

Associations were determined between Supera and the IVUS and non-IVUS groups, with examination of the clinical outcome variables: re-intervention, mortality, and amputation. T-tests or Chi-squares were used to compare continuous and categorical variables, respectively through the utility of IBM SPSS Statistical viewer. Associations were presented as Odds Ratios (O.R), Confidence Interval (C.I), p-value. A p≤0.05 was considered significant. All continuous variables were presented as mean ± standard deviation. Logistic regression models were also conducted in SPSS, using the clinical outcomes: re-intervention, mortality, and amputation as dependent variables and covariates as the variables that were considered significant in the univariate analysis.

## Results

A total of 236 SUPERA stents were used to treat 200 patients with lower limb claudication or critical limb ischemia. Ninety one of these patients received IVUS imaging, which was conducted in accordance with institution protocol. After angiographic review, all cases were universally agreed upon. 164 limbs were treated with a single Supera stent, while 36 limbs were treated with >1 Supera stents. In the case of the 36 limbs treated with >1 Supera stents, total stent length was used in order to determined compressed, nominal, or elongated.

Baseline and lesion characteristics were similar between both IVUS and non-IVUS treated cohorts. Vessel diameter was observed to be significantly larger in the IVUS subgroup (5.13mm vs 4.89mm, p<0.001), lesion length (117.47mm vs 90.64mm p<0.001), dissection (52.7% vs 22.0%, p<0.001) and calcification was also observed to be more frequent in the IVUS subgroup (Table 1 and 2).

**Table 1:**
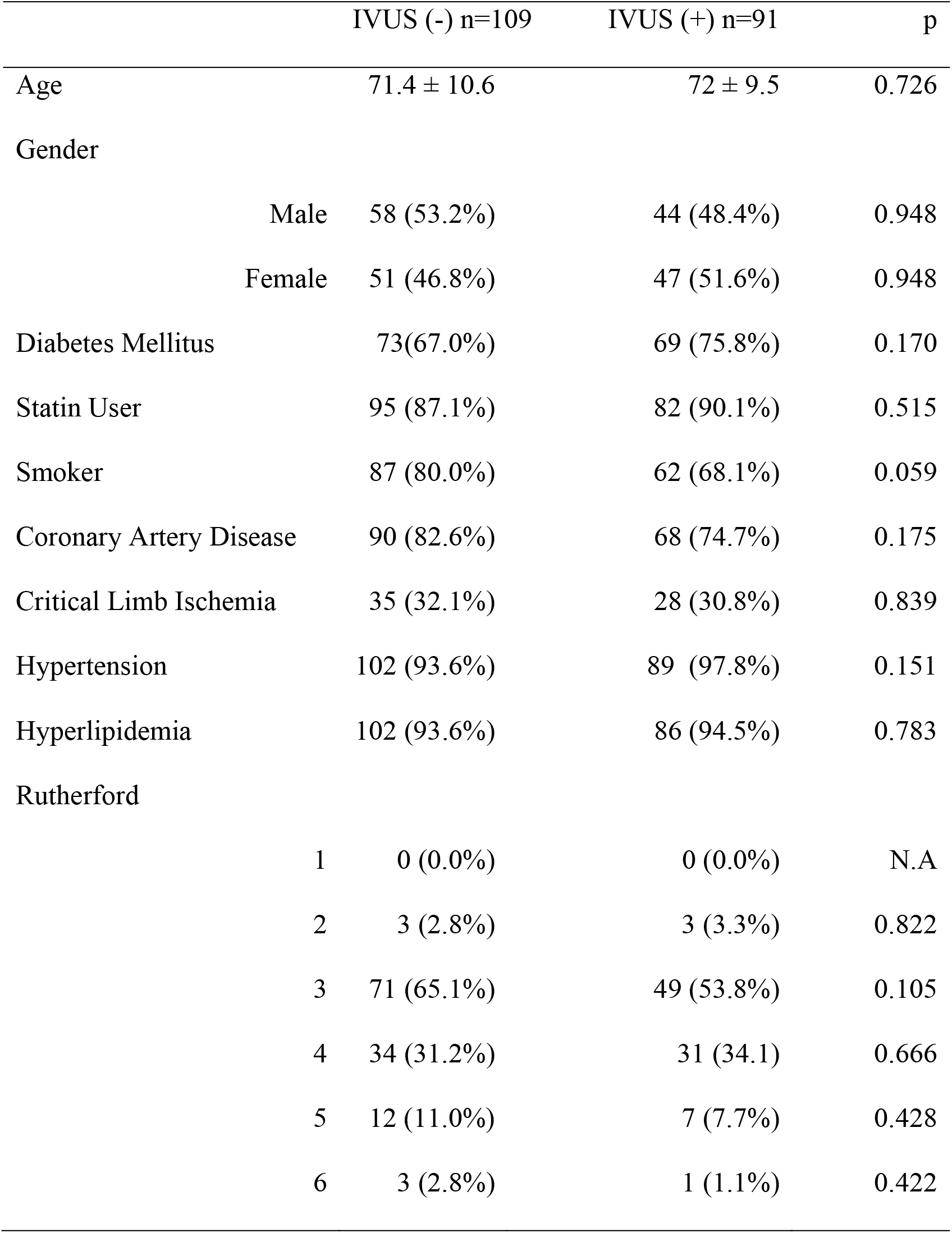
Baseline

**Table 2:** Clinical Parameters

| Table 2: Clinical Parameters | IVUS (-) n=109 | IVUS (+) n=91 | p |
| --- | --- | --- | --- |
| Reference Vessel Diameter |  |  |  |
| (mm) | 4.89 ± 0.79 | 5.13 ± 0.78 | <0.001 |
| 4 | 43 (39.4%) | 11 (12.1%) | <0.001 |
| 4.5 | 5 (4.6%) | 0 (0.0%) | N.A |
| 5 | 10 (9.2%) | 11 (12.1%) | 0.528 |
| 5.5 | 36 (33.0%) | 36 (39.6%) | 0.338 |
| 6 | 15 (13.8%) | 33 (36.3%) | <0.001 |
| Total Stent Length (mm) | 98.44 ± 36.44 | 120.3 ± 25.38 | 0.042 |
| 40 | 8 (7.3%) | 10 (11.0%) | 0.372 |
| 60 | 22 (20.2%) | 28 (30.8%) | 0.087 |
| 80 | 23 (21.1%) | 11 (12.1%) | 0.095 |
| 100 | 11 (10.1%) | 11 (12.1%) | 0.653 |
| 120 | 20 (18.3%) | 19 (20.9%) | 0.653 |
| 150 | 25 (22.9%) | 12 (13.2%) | 0.083 |
| Lesion Length (mm) | 90.64 ± 41.7 | 117.47 ± 49.3 | <0.001 |
| Average Vessel Runoff | 1.7 ± 1.0 | 1.7 ± 1.0 | 0.303 |
| Vessel Runoff (#) |  |  |  |
| 0 | 12 (11.0%) | 13 (14.3%) | 0.485 |
| 1 | 33 (30.3%) | 31 (34.1%) | 0.567 |
| 2 | 32 (29.4%) | 21 (23.1%) | 0.316 |
| 3 | 29 (26.6%) | 26 (28.6%) | 0.757 |
| Chronic Total Occlusion | 46 (42.2%) | 32 (35.2%) | 0.310 |
| Calcification | 14 (12.8%) | 35 (38.5%) | <0.001 |
| Dissection | 20 (22.0%) | 48 (52.7%) | <0.001 |
| Thrombus | 5 (4.6%) | 10 (11.0%) | 0.087 |
| Recoil | 4 (3.7%) | 1 (1.1%) | 0.246 |
| Perforation | 2 (1.8%) | 0 (0.0%) | 0.194 |

All vessel preparation devices were utilized at the discretion of the operator and included percutaneous transluminal angioplasty (PTA), drug coated balloons (DCB), atherectomies or thrombectomy (Table 3). There were no significant differences in the adjunctive methods between subgroups. There was a significant difference in the Supera stent size used within the IVUS + angiography cohort compared to angiography alone, with more 5.5mm (36/91 (40%) vs 36/109 (33%) p<0.05) and 6.0mm (33/91(36%) vs 15/109(14%), p<0.05). IVUS utility was associated with a lower CD-TLR: O.R = 0.482 [CI: 0.233,0.998] (p=0.047). There was no observed difference in lower extremity amputation between the two cohorts: O.R = 0.891 [CI:0.298, 2.670] (p=0.837).

**Table 3:** Adjunctive Therapies

| Table 3: Adjunctive Therapies | IVUS (-) n=109 | IVUS (+) n=91 | p |
| --- | --- | --- | --- |
| DCB | 48 (44.0%) | 38 (41.8%) | 0.746 |
| PTA | 61 (56.0%) | 54 (59.3%) | 0.630 |
| Thrombectomy | 3 (2.8%) | 0 (0.0%) | 0.910 |
| Laser Atherectomy | 1 (1.1%) | 0 (0.0%) | 0.951 |
| Directional Atherectomy | 86 (78.9%) | 75 (82.4%) | 0.532 |

IVUS was also associated with stent deployment status. Nominal deployment was achieved more frequently in the presence of IVUS utility: O.R = 2.674 [CI: 1.507,4.747], p<0.001), while elongated deployment was observed greater in the non-IVUS treated population (OR = 3.523 [CI: 1.709, 7.267], p<0.001) (Table 4). There was a greater rate of compressed deployment in the non-IVUS subgroup, however this was not statistically significant.

**Table 4:**
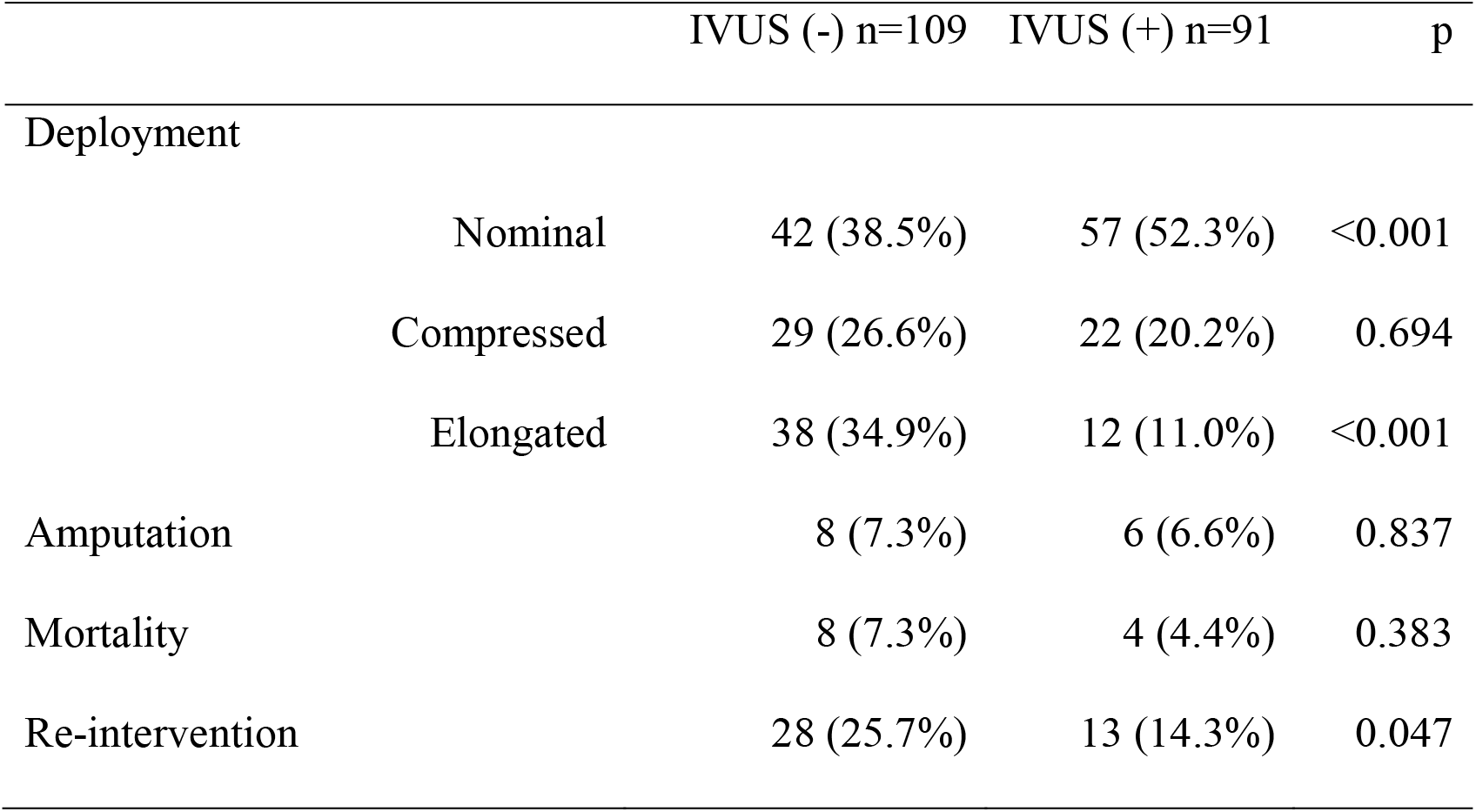
Outcomes

| Table 4: Outcomes |  | IVUS (-) n=109 | IVUS (+) n=91 | p |
| --- | --- | --- | --- | --- |
| Deployment |  |  |  |  |
|  | Nominal | 42 (38.5%) | 57 (52.3%) | <0.001 |
|  | Compressed | 29 (26.6%) | 22 (20.2%) | 0.694 |
|  | Elongated | 38 (34.9%) | 12 (11.0%) | <0.001 |
| Amputation |  | 8 (7.3%) | 6 (6.6%) | 0.837 |
| Mortality |  | 8 (7.3%) | 4 (4.4%) | 0.383 |
| Re-intervention |  | 28 (25.7%) | 13 (14.3%) | 0.047 |

The number needed to treat/number needed to harm (NNT/NNH), with the outcome of re-intervention, showed that on average, 8.3 patients would have to be treated with over not being treated with IVUS in order to have one additional patient not have an re-vascularization event.

## Discussion

Current findings of this study highlight the plausible benefit of adjunctive IVUS use with Supera stent utilization. Significant differences were observed in angiography + IVUS compared to angiography alone cohorts with regard to RVD (5.13mm vs 4.89mm, p<0.5), lesion length(117.47 vs 90.64, p<0.5), calcification (35 vs 14, p<0.001), and dissection (48 vs 20, p<0.001). Across the primary clinical outcomes, the group treated with IVUS-guidance and Supera proceeded to show a significantly lower rate of re-intervention in comparison to the Supera + angiography alone group at 12 months.

Across the board, IVUS has been well regarded for improving clinical outcomes within coronary and iliac lesions, and continues to grow prominence in the scope of peripheral intervention.(16-19) The addition of IVUS in peripheral vascular intervention, has been shown to reduce short term and long term major adverse limb events (MALE) and decreased binary restenosis in complete total occlusion (CTO).(20,21) A recent randomized controlled trial comparing angiography + IVUS to angiography alone in femoropopliteal interventions reported decreased binary restenosis when utilizing IVUS + angiography compared to angiography alone at 12 months; and observed a high rate of disagreement in imaging findings including RVD and lesion length, both of which were underestimated with angiography alone.(22) As endovascular therapies evolve the importance of precise imaging becomes heightened. In the case of DCB underestimating the RVD may result in poor apposition of DCB to vessel wall and ultimately suboptimal drug delivery.(22) Mal-deployment of SUPERA has been suggested as plausible explanation for the large variation of in CD-TLR across the literature with elongation carrying the greatest risk of restenosis compared to nominal deployment.(15) The addition of IVUS has been shown to affect medical decision making with regard to sizing of interventional device, while the goal of our study was not to assess change in management as a result of IVUS use, the authors did find significant differences in lesion characteristics including RVD, lesion length, calcification and dissection.(22) In addition to these findings the Supera devices used in IVUS + angiography arm were significantly larger than that of angiography alone arm and resulted in significantly more nominal deployments than angiography alone (52.3% vs 38.5%, p<0.001).

While Supera on its own has historically been effective at treating infra-inguinal superficial femoral artery lesions, the fact remains that sizing is a difficult variable to control.(15,23) In our study, the use of IVUS likely attributed to improved vessel prep, deployment, and ultimately more accurate sizing of Supera stent leading to the observed reduction in CD-TLR. Consistent use of IVUS may improve accurate vessel measurements and possibly reduce the error in stent deployment. These factors in combination could improve the long term clinical outcomes of endovascular procedures.

Due to the retrospective design of this study, only associations could be determined from the results. Another limitation was that mortality data was collected solely through EPIC records, since social security numbers were often unavailable, preventing cross verification with national death indexes. The present study also consists of a population from a heavily concentrated urban location, making out findings less representative of all populations.

## Conclusion

IVUS and angiography decreases clinically driven target lesion reintervention and increases nominal deployment compared to angiography alone in femoropopliteal lesions treated with interwoven bare metal nitinol stents.

### Perspectives

IVUS has been known to improve patency, protect from vascular adverse events, and reduce the need for reintervention. This study explores IVUS as an adjunctive imaging method in the management of femoropopliteal lesions with the Supera interwoven bare metal nitinol stents.IVUS with angiography shows improved nominal deployment and reduced CD-TLR when adjunctively used with Supera stents.

## Data Availability

Data is available on reasonable request.

## Abbreviations

PAD: Peripheral Artery Disease
CD-TLR: Clinically Driven Target Lesion Reintervention
IVUS: Intravascular Ultrasound
PTA: Percutaneous Transluminal Angioplasty
DCB: Drug Coated Balloon

## References

1. Shu J, Santulli G. Update on peripheral artery disease: Epidemiology and evidence-based facts. Atherosclerosis 2018;275:379–381.

2. Zhao HQ, Nikanorov A, Virmani R, Jones R, Pacheco E, Schwartz LB. Late stent expansion and neointimal proliferation of oversized Nitinol stents in peripheral arteries. Cardiovasc Intervent Radiol 2009;32:720–6.

3. Richard R S. The Importance of Accurate Femoropopliteal Artery Sizing in Endovascular Therapy. Endovascular Today 2012.

4. Fornell D. Understanding Intravascular Ultrasound (IVUS) Systems. Diagnostic and Interventional Cardiology 2009.

5. Iida O, Takahara M, Soga Y et al. Efficacy of intravascular ultrasound in femoropopliteal stenting for peripheral artery disease with TASC II class A to C lesions. J Endovasc Ther 2014;21:485–92.

6. Panaich SS, Arora S, Patel N et al. Intravascular Ultrasound in Lower Extremity Peripheral Vascular Interventions: Variation in Utilization and Impact on In-Hospital Outcomes From the Nationwide Inpatient Sample (2006-2011). J Endovasc Ther 2016;23:65–75.

7. Krishnan P, Tarricone A, Gee A et al. Analysis of Interwoven Nitinol Stenting for the Treatment of Critical Limb Ischemia: Outcomes From an Average 3-Year Follow-up Period. Angiology 2022;73:407–412.

8. Hiramoto JS, Teraa M, de Borst GJ, Conte MS. Interventions for lower extremity peripheral artery disease. Nat Rev Cardiol 2018;15:332–350.

9. Meng FC, Chen PL, Lee CY, Shih CC, Chen IM. Real-World Comparison of Drug-Eluting and Bare-Metal Stents in Superficial Femoral Artery Occlusive Disease with Trans-Atlantic Intersociety Consensus B Lesions: A 2-Year, Single-Institute Study. Acta Cardiol Sin 2018;34:130–136.

10. Ruiz CF, San Norberto García E, Domingos LF et al. supera Stent in Popliteal Artery, 2 Years Follow-up Results. European Journal of Vascular and Endovascular Surgery 2019;58:e455.

11. Werner M, Paetzold A, Banning-Eichenseer U et al. Treatment of complex atherosclerotic femoropopliteal artery disease with a self-expanding interwoven nitinol stent: midterm results from the Leipzig SUPERA 500 registry. EuroIntervention 2014;10:861–8.

12. Garcia L, Jaff MR, Metzger C et al. Wire-Interwoven Nitinol Stent Outcome in the Superficial Femoral and Proximal Popliteal Arteries: Twelve-Month Results of the SUPERB Trial. Circ Cardiovasc Interv 2015;8.

13. de Boer SW, van den Heuvel DAF, de Vries-Werson DAB et al. Short-term Results of the RAPID Randomized Trial of the Legflow Paclitaxel-Eluting Balloon With Supera Stenting vs Supera Stenting Alone for the Treatment of Intermediate and Long Superficial Femoral Artery Lesions. J Endovasc Ther 2017;24:783–792.

14. San Norberto EM, Flota CM, Fidalgo-Domingos L, Taylor JH, Vaquero C. Real-World Results of Supera Stent Implantation for Popliteal Artery Atherosclerotic Lesions: 3-Year Outcome. Ann Vasc Surg 2020;62:397–405.

15. Garcia LA, Rosenfield KR, Metzger CD et al. SUPERB final 3-year outcomes using interwoven nitinol biomimetic supera stent. Catheterization and Cardiovascular Interventions 2017;89:1259–1267.

16. Buckley CJ, Arko FR, Lee S et al. Intravascular ultrasound scanning improves long-term patency of iliac lesions treated with balloon angioplasty and primary stenting. J Vasc Surg 2002;35:316–23.

17. Kim BK, Shin DH, Hong MK et al. Clinical Impact of Intravascular Ultrasound-Guided Chronic Total Occlusion Intervention With Zotarolimus-Eluting Versus Biolimus-Eluting Stent Implantation: Randomized Study. Circ Cardiovasc Interv 2015;8:e002592.

18. Liu XM, Yang ZM, Liu XK et al. Intravascular ultrasound-guided drug-eluting stent implantation for patients with unprotected left main coronary artery lesions: A single-center randomized trial. Anatol J Cardiol 2019;21:83–90.

19. Tan Q, Wang Q, Liu D, Zhang S, Zhang Y, Li Y. Intravascular ultrasound-guided unprotected left main coronary artery stenting in the elderly. Saudi Med J 2015;36:549–53.

20. Divakaran S, Parikh SA, Hawkins BM et al. Temporal Trends, Practice Variation, and Associated Outcomes With IVUS Use During Peripheral Arterial Intervention. JACC Cardiovasc Interv 2022;15:2080–2090.

21. Tsujimura T, Iida O, Takahara M et al. Clinical Impact of Intravascular Ultrasound–Guided Fluoropolymer-Based Drug-Eluting Stent Implantation for Femoropopliteal Lesions. JACC: Cardiovascular Interventions 2022;15:1569–1578.

22. Allan RB, Puckridge PJ, Spark JI, Delaney CL. The Impact of Intravascular Ultrasound on Femoropopliteal Artery Endovascular Interventions: A Randomized Controlled Trial. JACC Cardiovasc Interv 2022;15:536–546.

23. Palena LM, Diaz-Sandoval LJ, Sultato E et al. Feasibility and 1-Year outcomes of subintimal revascularization with supera(^®^) stenting of long femoropopliteal occlusions in critical limb ischemia: The “Supersub” Study. Catheter Cardiovasc Interv 2017;89:910–920.

